# Plasma acellular transcriptome contains Parkinson’s disease signatures that can inform clinical diagnosis

**DOI:** 10.1101/2024.10.18.24315717

**Authors:** Aleksandra Beric, Alejandro Cisterna-García, Charissa Martin, Ravindra Kumar, Isabel Alfradique-Dunham, Kevin Boyer, Ibrahim Olabayode Saliu, Shinnosuke Yamada, Jessie Sanford, Daniel Western, Menghan Liu, Ignacio Alvarez, Joel S. Perlmutter, Scott A. Norris, Pau Pastor, Guoyan Zhao, Juan Botia, Laura Ibanez

## Abstract

We aimed to identify plasma cell-free transcripts (cfRNA) associated with Parkinson’s disease (PD) that also have a high predictive value to differentiate PD from healthy controls. Leveraging two independent populations from two different movement disorder centers we identified 2,188 differentially expressed cfRNAs after meta-analysis. The identified transcripts were enriched in PD relevant pathways, such as PD (p=9.26×10^-4^), ubiquitin-mediated proteolysis (p=7.41×10^-5^) and endocytosis (p=4.21×10^-6^). Utilizing in-house and publicly available brain, whole blood, and acellular plasma transcriptomic and proteomic PD datasets, we found significant overlap across dysregulated biological species in the different tissues and the different biological layers. We developed three predictive models containing increasing number of transcripts that can distinguish PD from healthy control with an area under the ROC Curve (AUC) ≥0.85. Finally, we showed that several of the predictive transcripts significantly correlate with symptom severity measured by UPDRS-III. Overall, we have demonstrated that cfRNA contains pathological signatures and has the potential to be utilized as biomarker to aid in PD diagnostics and monitoring.

## INTRODUCTION

Parkinson’s disease (PD) is a slowly progressing, complex neurodegenerative disorder, with higher prevalence in males.^1,2^ It is one of the most common neurodegenerative diseases (NDDs), second only to Alzheimer’s disease (AD).^3,4^ As with other NDDs, the greatest risk factor for PD development is age, with incidence peaking after 80 years of age, with contributions from environmental and genetic factors.^2^ PD is characterized pathologically by formation of Lewy bodies (LBs) and early death of dopaminergic neurons, resulting in a typical clinical presentation including bradykinesia, rest tremor and rigidity. Other clinical hallmarks of PD include a number of non-motor symptoms, such as sleep, gastrointestinal and olfactory disorders, which may precede motor disorders by over a decade.^2,4,5^ At the molecular level, LBs are primarily comprised of misfolded α-synuclein, which can spread between the cells, serving as a template for further α-synuclein misfolding.^5^

While PD diagnoses largely depend on patient history and physical examination, no currently available tests enable definitive diagnosis of PD in the early stages.^2,4^ Instead, definitive diagnostics presently depends on neuropathological analyses upon death, typically occurring many years after disease onset.^1,2,4^ Several imaging methods can aid in confirm nigrostriatal deficits that occur in PD, but are not diagnostic.^6–9^ Dopamine transporter single-photon emission computed tomography (DaT SPECT) can detect cell loss in PD patients^4,6^, while positron emission tomography (PET) scan can point to early signs of dopaminergic neuron damage.^4,7,10,11^ Conversely, magnetic resonance imaging (MRI) methods provide modest benefit for diagnosis of PD.^4,6,7,10^ In addition to imaging, the field has strived to develop PD-specific cerebrospinal fluid (CSF) biomarkers, independent of clinical representation of the disease.^12^ CSF levels of α-synuclein have been the focal point of a number of studies. α-synuclein seed amplification assays (SAA) have shown a lot of promise, with the ability to differentiate between PD and healthy controls.^13,14^ However, results have been variable possibly due to clinical heterogeneity, cross-contamination with blood, or experimental differences, requiring further validation prior to clinical implementation. Lysosomal enzymes and neurofilament light chain emerged as candidates for biomarker panels, though they also require further investigation.^12,15,16^ Another unmet need is the differential diagnosis of PD from other NDDs. Though it is a very common neurodegenerative disorder, PD is misdiagnosed in clinical practice with error rates reported to range from 15% to 24%.^4^ The prevailing reason for disagreement between clinical and neuropathological diagnoses is the heterogeneity of parkinsonism with non-PD pathologies, including multiple system atrophy (MSA) and progressive supranuclear palsy (PSP).^17^ Indeed, even established PD cases are greatly heterogeneous in the age of onset, rate of progression as well as clinical presentation, which led to the establishment of several PD subtypes.^4,6^

While most blood-based biomarker studies focus on measuring levels of various proteins, circulating nucleic acids have found their place in clinical practice. Analyses of cell-free DNA (cfDNA) have revolutionized the field of obstetrics and antenatal testing by allowing the identification of fetal aneuploidies in a sample of mother’s blood, thus reducing test-related risk of miscarriage.^18–20^ cfDNA has been utilized as a biomarker for cancer^20–22^, metabolic disorders^20^ and a way to assess the health of donor organs in a recipient’s body upon organ transplantation.^20,23^ In addition to cfDNA, cell-free RNA (cfRNA) can also be captured from plasma and provides a temporal snapshot of cellular processes throughout the body, as it is released from cells as part of normal cell death.^24^ Numerous studies are investigating the potential of using cfRNAs as biomarkers for prenatal testing^19^, cancer^21,25–28^ and AD.^29–33^ Furthermore, a recent study provided evidence of circulating micro RNAs being involved in the regulation of PD-associated genes^34^, adding support to the utility of non-protein biomarkers in the PD field.

In this study, we used plasma cfRNA from two independent populations of PD participants to capture transcriptional changes caused by PD pathology. We biologically contextualized our findings via pathway analyses and multiomic data integration by accessing whole blood and brain transcriptomic datasets and plasma and CSF proteomic datasets. Then we leveraged those to build a predictive model that could accurately predict PD using a limited number of transcripts, with the potential for translation into clinical practice. We further evaluated the capabilities of the best performing models to discriminate between PD and AD, dementia with Lewy bodies (DLB) and frontotemporal dementia (FTD) to ensure that captured changes were specific to PD pathobiology.

## METHODS

### Study design

We analyzed acellular RNAseq data from two independent movement disorder clinical cohorts, Hospital Universitari Mutua Terrassa (HUMT) in Barcelona, Spain, and Washington University in Saint Louis School of Medicine (WUSM) in Saint Louis, US. We performed differential expression (DE) analyses in HUMT (206 participants) and WUSM (175 participants) cohorts separately, followed by meta-analysis. Transcripts with Benjamini-Hochberg corrected p-values below 0.05 in the meta-analysis were considered DE. To understand the biological significance of the DE transcripts, we performed pathway analyses and leveraged in-house and publicly available datasets to contextualize the expression of the identified transcripts in whole blood and brain, as well as the accumulation of corresponding proteins in plasma and CSF. To assess the diagnostic capabilities of cfRNAs, we developed several predictive models, focusing on the DE transcripts. We then employed a third independent dataset, consisting of participants with dementia with DLB, AD, and FTD, to test the specificity of the predictive models for PD. AD, DLB, and FTD participants were diagnosed according to the clinical criteria contained in the Uniform Data Set (UDS), the standard set of clinical data collected in all participants enrolled in any of the 37-federally funded ADRCs. Finally, utilizing the information available about participants’ motor symptom severity, measured by Unified Parkinson’s Disease Rating Scale Part III (UPDRS-III), and cognitive status measured by The Montreal Cognitive Assessment (MoCA), we assessed the clinical relevance of transcripts included in the predictive models. Research in this study was conducted in accordance with recommended protocols. Written informed consent was obtained from all participants or their family members. The Washington University in Saint Louis Institutional Review Board approved the study (IRB ID 201701124 and 202004010).

### Study Participants

We obtained plasma samples from two independent cohorts, HUMT and WUSM. HUMT cohort included a total of 206 plasma samples (87 PD participants and 119 healthy controls), while the WUSM cohort included 175 samples (94 PD participants and 81 healthy controls). Given the different geographical location and standards of care, the two cohorts show some differences (Table 1). They are comparable in proportion of female participants (HUMT 47.57%, WUSM 48.57%; p=0.93). The HUMT population shows a lower mean age (68.26±8.39) compared to the WUSM population (72.97±6.78; p= 3.30×10^-9^). Differences are also observed in motor symptom severity as measured by the UPDRS-III scale. Participants in the WUSM population displayed greater UPDRS-III (26.03±9.10), compared to HUMT participants (18.69±8.99; p=6.39×10^-4^). Finally, dementia was assessed only in the WUSM participants, who presented with mild or no signs of dementia (25.32±4.22), as measured by the MoCA scale. Similarly, no therapeutic information was available for the HUMT dataset.

**Table 1.** Summary demographics of the two populations included in the main analyses.

|  | Hospital Universitari Mutua Terrassa<br>(Barcelona, Spain) |  | Washington University School of<br>Medicine (Saint Louis, US) |  |
| --- | --- | --- | --- | --- |
|  | Healthy Controls | Parkinson's<br>Disease | Healthy Controls | Parkinson's<br>Disease |
| **Participants (N)** | 119 | 87 | 81 | 94 |
| **Mean age (IQR)** | 67.78 (61.00-74.00) | 68.91 (65.00-74.00) | 74.07 (69.00-78.00) | 72.02 (67.00-77.00) |
| **Female (N, %)** | 63 (52.94%) | 35 (43.21%) | 49 (60.49%) | 36 (38.29%) |
| **Mean UPDRS-III**<br>**(IQR)** | - | 18.70 (14.00-22.00) | - | 26.03 (20.69-30.00) |
| **Mean MoCA (IQR)** | - | - | - | 25.32 (24.00-28.00) |

N=Sample size; IQR=Interquartile Range; UPDRS=Unified Parkinson's Disease Rating Scale; MoCA=Montreal Cognitive Assessment; US=United States

### RNA Extraction and Sequencing

Whole blood samples were collected from all participants. Within 20 min of collection, blood samples were centrifuged for 10 min at 1500rpm to obtain plasma and subsequently stored at - 80°C, as previously described.^35^ Plasma samples were thawed on ice and centrifuged for 5 min at 2000 rpm prior to RNA extraction to remove any cells present and avoid cellular RNA contamination. Total plasma cfRNA was extracted from 0.5 mL of plasma using the Maxwell RSC miRNA from plasma or serum kit (Ambion) and ribodepleted (NEBNext rRNA Depletion Kit). After total RNA quantification, libraries were generated using the NEBNext Ultra II Directional RNA Library Prep Kit for Illumina (New England Biolabs) using 1ng of RNA as input. Libraries were cleaned for adapter dimers prior to sequencing. We targeted 40 million 100 base pair single-end reads for each sample using an Illumina NovaSeq 6000.

### Data Processing and Quality Control

FastQC (v0.11.7)^36^ was used to evaluate the sequencing quality of each sample. Reads were aligned to the human reference genome GRCh38 using STAR (v2.7.1a).^37^ The quality of sequences and alignments was assessed with PICARD (v2.26)^38^ and SAMtools^39^, and transcripts quantified using Salmon (v0.11.3).^40^ Quality control measures were gathered via MultiQC (v1.9)^41^ followed by stringent quality control (QC). Briefly, all genes with less than ten reads in 90% or more of the individuals were removed. Subsequently, transcriptome Principal Component Analysis (PCA) were performed and screened for correlation with technical and methodological variables to detect potential biases. A strong correlation was observed with total reads and coding bases; thus, all samples with less than 10% of coding bases and less than 1,000,000 total reads were removed. Outlier samples identified via transcriptome PCA, defined as samples whose first two principal component values deviated more than three standard deviations from mean values of either of the respective principal components, were also removed.

Despite following the same protocol for data generation, processing, and QC, the two datasets included in the present study (HUMT and WUSM) were sequenced at different timepoints. Each dataset underwent QC separately. ComBat_seq function from the sva package^42^ was used to adjust for technical variation within each dataset. Batch effect correction was followed by PCA and removal of any additional outliers. As previously described, there is RNA degradation associated with plasma long-term storage (up to 20 years).^33^ Consequently, we addressed degradation using DESeq2 (v1.22.2)^43^ to find transcripts associated with storage time in control participants as previously reported.^33^ All transcripts nominally (p<0.05) associated with storage time were removed from the analyses from both HUMT (n=486 transcripts) and WUSM (n=221 transcripts) datasets. Further, due to prevalence of PD therapies and lack of medication data for HUMT population, transcripts associated with PD-medication usage were identified in the WUSM dataset using DESeq2^43^ and any nominally significant transcripts (p<0.05) were removed from further analyses from both datasets (n=630 transcripts). Overall, 27832 transcripts passed QC and were included used in subsequent analyses.

### Differential Expression Analyses and Pathway Analyses

Differential expression (DE) analyses were performed using DESeq2^43^ in HUMT and WUSM data separately, followed by meta-analysis using metaRNASeq.^44^ All analyses were adjusted by sex and age at blood draw. Benjamini-Hochberg correction (FDR) was used to correct for multiple testing, considering meta-analysis FDR p-values lower than 0.05 as significant. No effect size (log_2_ fold change) value cut-off was applied. Pathway enrichment analyses were carried out using clusterProfiler^45^ to functionally characterize the identified transcripts and FDR p-values below 0.05 were regarded as significant.

### Multi-omic data integration

To biologically contextualize our findings, we cross-checked the results from the cfRNA DE meta-analysis with: (i) in-house brain bulk RNAseq, (ii) publicly available, whole blood bulk RNAseq^46^, (iii) publicly available plasma and (iv) CSF proteomic data^46^, and (v) in house and (vi) publicly available brain single-cell RNAseq data^47,48^ including substantia nigra scRNAseq (Figure 1). Following differential expression or accumulation analyses in each of the RNAseq or proteomic datasets, we identified the overlap between nominally significant findings and the DE plasma acellular transcripts and each accessed dataset. The significance of each overlap was tested using hypergeometric test (phyper function in R), p-values lower than 0.05 considered significant.

**Figure 1.**
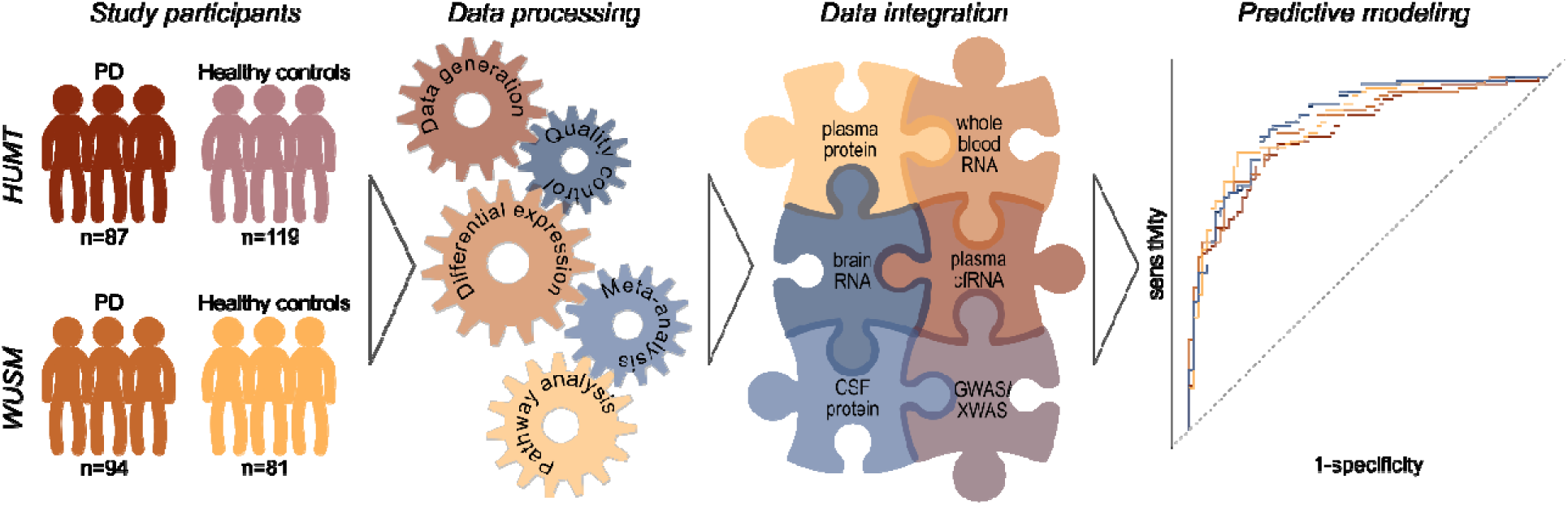
Study Design Infographic depicting the study participants from two independent cohorts, summary of the experimental approach including data processing and analyses, biological contextualization of the results via multiomic data integration with available independent datasets, and leverage of results to build predictive models.

Additionally, to investigate whether any of the DE transcripts mapped to PD associated loci identified by genome wide association analyses (GWAS), we leveraged the information available in the PD GWAS locus browse and in the latest multi-ancestry genome-wide association study.^49,50^ We browsed the PD GWAS locus browser and recorded the scores that rank the genes based on the amount of supporting evidence compiled in the PD GWAS locus browser.^50^ Next, to compare our results to the multi-ancestry GWAS, we converted locations of all lead SNPs from hg19 version of the human genome to the respective locations in hg38 version of the human genome and then found coordinates 500kb upstream and downstream from each SNP.^49^ All regions that overlapped after the coordinate conversion were collapsed into 69 non-overlapping genomic regions. Subsequently, collapsed PD-associated genomic regions were overlapped with genomic start and end coordinates of the identified DE genes using bedtools intersect from BEDTools tool suite.^51^

### Predictive Models Construction and Evaluation

To build and evaluate predictive models we used the previously published in-house pipeline.^33^ In brief, glmnet (v4.1.6)^52^ was leveraged to produce a suitable classifier to identify PD cases based on plasma acellular gene expression. HUMT was used as the training and WUSM as the testing dataset. After ComBat_seq^42^ regression, transcript counts were further scaled between the two datasets by computing the z-score using the mean and standard deviation. With the transcripts that were significantly DE in the meta-analysis after multiple test correction (FDR<0.05), we calculated Kullback-Leibler divergence (KLD) between the training (HUMT) and the testing (WUSM) dataset for each transcript using R package entropy v1.3.1.^53^ A hundred L2 regularization linear models were trained with increasing number of transcripts, ranging in KLD value from 0.01 to 1 in increments of 0.01. The Area Under the Receiver Operating Characteristic (ROC) Curve (AUC) value was computed for all models in the training dataset.

### Specificity Analyses

Performance of the best predictive models was evaluated in 44 AD, 16 FTD and 17 DLB cases from a publicly available dataset from the Knight-ADRC^33,54^, as well as twelve DLB participants available in the HUMT dataset. Knight-ADRC data has been generated and processed as described above for the HUMT and WUSM datasets. Transcript counts were scaled by computing the z-score using the mean and standard deviation. Then, risk score for each individual was calculated using the previously defined predictive models. Scores higher than 0.50 were considered cases. ROC curve was computed by comparing the predicted to the actual disease status for each compared group. We assessed the ability of the predictive models to differentiate between AD, DLB or FTD and healthy controls, as well as between AD, DLB or FTD and PD. Additionally, we evaluated if the addition of the APOE genotype to the cfRNA predictor improved the model performance in differentiating between PD and AD, as APOE is a crucial genetic risk factor for AD. APOE genotype data was available for AD and WUSM PD participants. To discern whether the effect of APOE was captured by the predictor, we included the APOE genotype in the model coded by two variables representing the number of ε2 alleles and ε4 alleles.

### Evaluation of predictive models’ clinical relevance

To explore the relationship of selected transcripts with PD clinical manifestations, we calculated Spearman correlations between normalized and age and sex adjusted transcript counts with UPDRS-III or MoCA scores. We did it for those PD participants with available data. UPDRS-III information was available for 76 participants from HUMT and 27 participants in the WUSM population. To maximize our sample size and statistical power for this exploratory analysis, the two populations were combined for correlation with UPDRS-III. MoCA scores were not available for the HUMT population, thus correlation to MoCA analysis was carried out only in the WUSM population (n=28). Correlations were considered significant if p-value was lower than 0.05.

## RESULTS

### Acellular transcriptomic patterns translate to changes of plasma protein abundances

We leveraged two independent movement disorder clinics (Hospital Universitari MutuaTerrassa (HUMT), and Washington University School of Medicine (WUSM)) with a total of 181 PD participants (*n*_HUMT_=87; *n*_WUSM_=94) and 200 healthy control participants (*n*_HUMT_=119; *n*_WUSM_=81) (Figure 1, Table 1). All PD participants had a clinical diagnosis of PD at the time of sample collection.^55^ After stringent quality control (QC), we performed differential expression (DE) analyses comparing PD to healthy control participants using DESeq2^43^, followed by integration of HUMT and WUSM results through meta-analysis. We performed meta-analyses on 6,496 transcripts which had same direction of effect in both HUMT and WUSM populations, and identified 2,188 DE transcripts, 1,101 of which were upregulated and 1,087 downregulated (Supplementary Figure 1, Supplementary Table 1). To evaluate the biological relevance of the DE transcripts, we performed Kyoto Encyclopedia of Genes and Genomes (KEGG) pathway enrichment analyses. We found significant enrichment in PD (p=9.26×10^-4^), endocytosis (p=4.21×10^-6^) and ubiquitin mediated proteolysis (p=7.41×10^-5^), along with other nervous system diseases such as Huntington’s disease (HD; p=6.35×10^-4^) and amyotrophic lateral sclerosis (ALS; p=6.10×10^-4^; Figure 3A, Supplementary Table 2). Additionally, we preformed Gene Ontology (GO) enrichment analyses and found enrichment in the cellular components primary lysosome (p=4.19×10^-3^) and ubiquitin ligase complex (p=2.12×10^-3^), and biological processes such as late endosome to lysosome transport (p=8.18×10^-4^) and exocytosis (p=9.48×10^-6^; Supplementary Table 3).

To further replicate our results, we used publicly available whole blood RNAseq data^56^ from the Parkinson’s Progression Markers Initiative (PPMI)^46^ and Parkinson’s Disease Biomarkers Program (PDBP)^57^ and found 424 of the 2,188 DE transcripts identified in plasma were also DE in whole blood^56^ (Figure 2A), which represents a significant overlap (p=4.65×10^-60^). These 424 transcripts were enriched in ubiquitin mediated proteolysis (p=8.96×10^-5^; Supplementary Table 2). Additionally, we investigated if the abundances of the proteins encoded by the identified RNAs were also significantly different. We leveraged plasma proteomic data from PPMI generated with Olink. We found a significant overlap (p=5.00×10^-3^) of twelve differentially accumulated proteins out of the 2188 transcripts DE in plasma (Figure 2A). Further, six of the twelve proteins have the same direction of effect as the respective mRNAs from plasma (Supplementary Table 1). Looking back to the pathway analysis, we find that the twelve proteins were present in the immune response pathways. Two of the twelve acellular transcripts that are differentially accumulated in plasma, at both transcript and protein level, are also DE in whole blood (Figure 2A, Supplementary Table 1).

**Figure 2.**
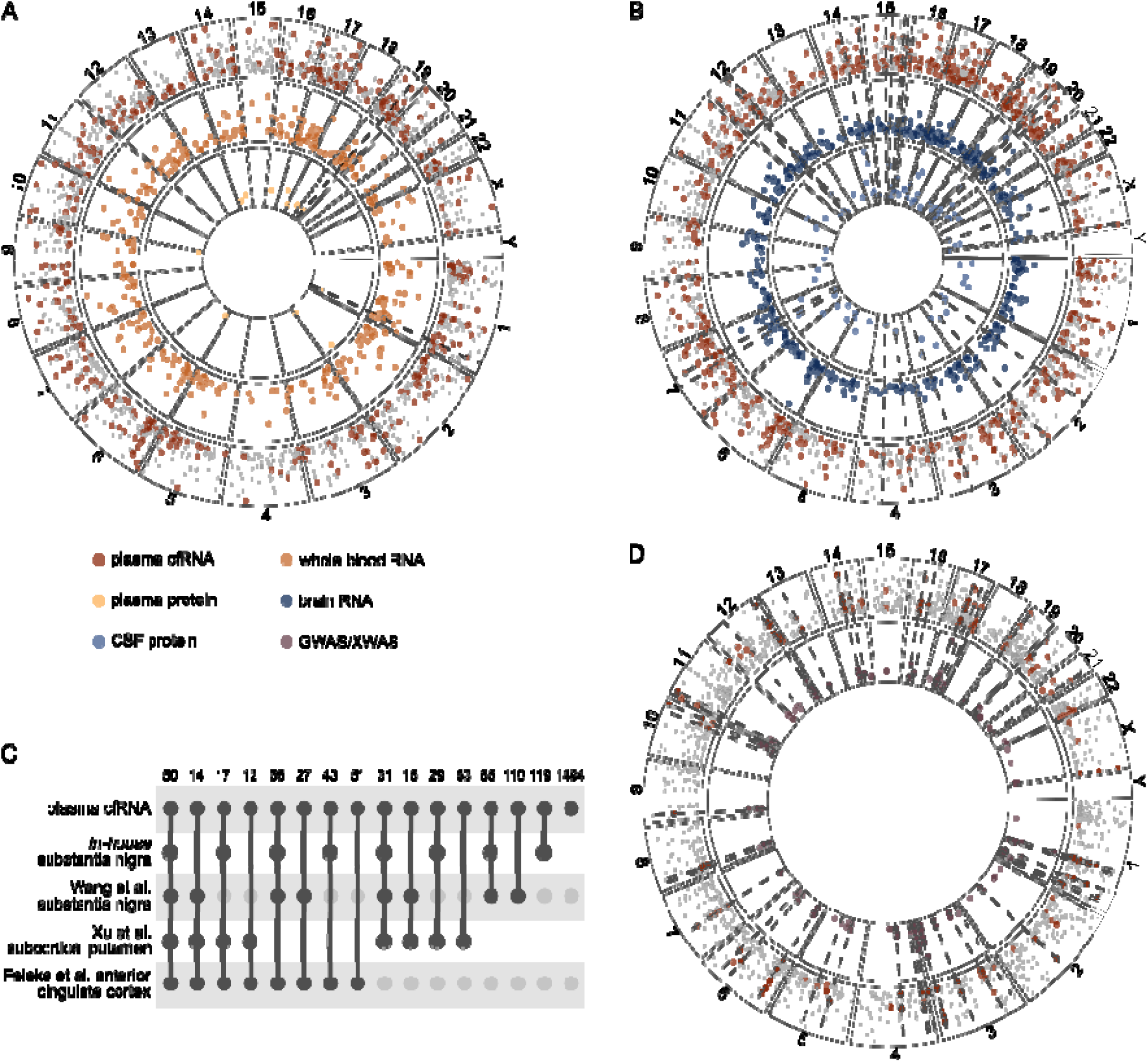
Multiomic integration summary. Integration between differential expression analysis of cfRNA (maroon) with **A**. whole blood RNAseq (orange) and plasma proteomics (light yellow); **B**. brain RNAseq (dark blue) and CSF proteomics (light blue); **C**. single nuclei datasets; and **D**. GWAS data (rosy-brown). The y-axes in panels A, B and D represent -log_10_ of differential expression p-value, while x-axes represent genomic coordinates of the respective transcript/gene. Dotted lines symbolize the same transcript/gene or its protein product that are significantly differentially expressed across the multiple *<u>omic</u>* layers displayed in each respective panel, A, B or D.

### Expression patterns in plasma follow those observed in brain

We compared our findings to an in-house brain RNAseq dataset^54^ to test whether the changes we capture in plasma might have their origin in pathological changes taking place in the brain. Out of the 2,188 transcripts, we found 537 transcripts that were DE in both plasma and brain (p=4.22×10^-105^; Figure 2B, Supplementary Table 1), 278 of which were upregulated and 259 downregulated. Of the 537 transcripts, 286 (142 upregulated and 144 downregulated) have the same direction of effect in plasma and brain. Transcripts that were DE in both plasma and brain were enriched in neurodegenerative diseases including PD (p=7.86×10^-7^), HD (p=6.27×10^-8^), and AD (p=2.16×10^-6^), as well as endocytosis (p=8.91×10^-4^) and ubiquitin mediated proteolysi (p=2.47×10^-3^; Supplementary Table 2).

Next, we compared our DE transcripts with differentially expressed genes derived from three independent single-nucleus RNA-seq datasets from individuals with PD and controls: i) subcortical putamen tissue^48^, ii) anterior cingulate cortex^47^, and iii) *in-house* and publicly available^58^ substantia nigra. Out of the 2,188 transcripts DE in plasma, 222 were also DE in at least one cell type in the subcortical putamen (Figure 2C, Supplementary Table 4). The greatest overlap between plasma and subcortical putamen was in transcripts expressed by ependymal cells (a type of glial cells), with 162 shared DE transcripts, followed by 41 transcripts in GRIK3-enriched neuronal cells and eleven transcripts in astrocytes (Supplementary Table 4). Transcripts DE in both plasma and subcortical putamen were nominally enriched in HD (p=3.51×10^-3^) and general pathways of neurodegeneration (p=0.03; Supplementary Table 2). Next, we found 77 DE transcripts shared between plasma and the anterior cingulate cortex. The greatest overlap between plasma and anterior cingulate cortex was in neuronal cells (47 transcripts), followed by myeloid cells (27 transcripts), while only three transcripts were shared between plasma cfRNA and cortical astrocytes (Supplementary Table 5). These transcripts were enriched in PD (p=7.59×10^-4^), as well as pathways of neurodegeneration (p=1.35×10^-3^, Supplementary Table 2). Of the 222 transcripts shared between plasma and subcortical putamen and 77 transcripts shared between plasma and anterior cingulate cortex, 35 were DE in both subcortical putamen and anterior cingulate cortex (Figure 2C, Supplementary Tables 4 and 5).

*In-house* substantia nigra had 411 DE transcripts in common with plasma (Figure 2C, Supplementary Table 6). These transcripts were enriched in PD (p=3.35×10^-4^), endocytosis (p=4.26×10^-5^) and ubiquitin mediated proteolysis (p=6.92×10^-4^; Supplementary Table 2). Similar to subcortical putamen, ependymal cells from the *in-house* substantia nigra dataset showed the greatest overlap with plasma cfRNA transcripts (245 transcripts; Supplementary Table 6). In parallel, 370 of the 2,188 DE cfRNA were DE in the public substantia nigra dataset^58^ (Figure 2C, Supplementary Table 7). They were enriched in ubiquitin mediated proteolysis (p=6.06×10^-5^), dopaminergic synapse (p=8.19×10^-3^) and general pathways of neurodegeneration (p=9.10×10^-3^; Supplementary Table 2). Furthermore, 229 plasma DE transcripts were shared with both the in-house and the public substantia nigra. Additionally, 72 of those 229 were in the same direction of effect in the two substantia nigra datasets.

Finally, using publicly available CSF proteomic data generated with Somalogic (PPMI)^46^, we tested whether any of the DE transcripts correspond to differentially abundant CSF proteins. We uncovered a significant overlap (p=4.90×10^-6^) of 89 acellular plasma transcripts and their respective protein products in CSF (Figure 2B), which were enriched in endocytosis (p=8.10×10^-7^) and nominally enriched in HD (p=1.83×10^-2^), and general pathways of neurodegeneration (p=4.40×10^-2^; Supplementary Table 2). Of the 89 transcripts whose protein products were differentially accumulated in CSF, 24 are also DE in bulk brain RNAseq (Supplementary Table 1).

### Plasma differentially expressed transcripts originate from PD-risk loci

We then investigated if any of the transcripts DE in plasma was encoded in PD risk loci. We detected a significant overlap of 190 transcripts (p=2.88×10^-272^) that mapped to PD-associated GWAS loci^49^ corresponding to 69 non-overlapping genomic regions (Figure 2D, Supplementary Table 8). We found an average of 3 (±2) overlapping transcripts per genomic region, 27 regions overlapping with only one of the identified transcripts, and 14 regions overlapping five to nine transcripts each. Further, we found that six DE transcripts overlapped (p=1.47×10^-8^) PD-associated loci on the X chromosome^59^ (Supplementary Table 8). Next, we compared the nominated genes in the PD GWAS locus browser^50^ with the DE transcripts identified in our analyses to assess if there is an agreement with the nominated gene. We found varying levels of support for 121 of the 190 autosomal genes, with an average score of 4.42±1.88 (Supplementary Table 8). Genes located on the X chromosome cannot be evaluated due to the unavailability of sex chromosome data in the GWAS browser. We checked if any of the 121 genes were DE in whole blood or brain RNAseq, or if their protein products were differentially accumulated in plasma or CSF. We found 26 of the 121 genes DE In whole blood and 24 of the 121 DE in brain (Supplementary Tables 1 and 8). Similarly, we observed that corresponding protein products of two of the 121 genes (ITGAM and PARP1) were differentially accumulated in plasma, and four (GCH1, HEXIM2, LGALS3 and UFC1) differentially accumulated in CSF (Supplementary Tables 1 and 8).

### cfRNA captures transcriptomic signatures corresponding to Parkinson’s disease

We wanted to assess if cfRNA changes can be leveraged to build predictive models. To utilize the two independent RNAseq data sets for the development of predictive models, we employed an approach similar to that used previously focusing only on the 2,188 DE transcripts.^33^ HUMT was used as training and WUSM as testing. We generated a total of 100 predictive models corresponding to 100 KLD threshold values (increments of 0.01). Based on the balance between ROC-AUC and number of transcripts included in the model we selected three transcript subsets for further follow up (Supplementary Figure 2). Of note, each larger subset is inclusive of the smaller ones. The three selected subsets contained 26, 87 and 191 transcripts, with ROC-AUC values of 0.86, 0.87 and 0.88 in the testing data set (Figure 3B, Supplementary Table 9), respectively.

**Figure 3.**
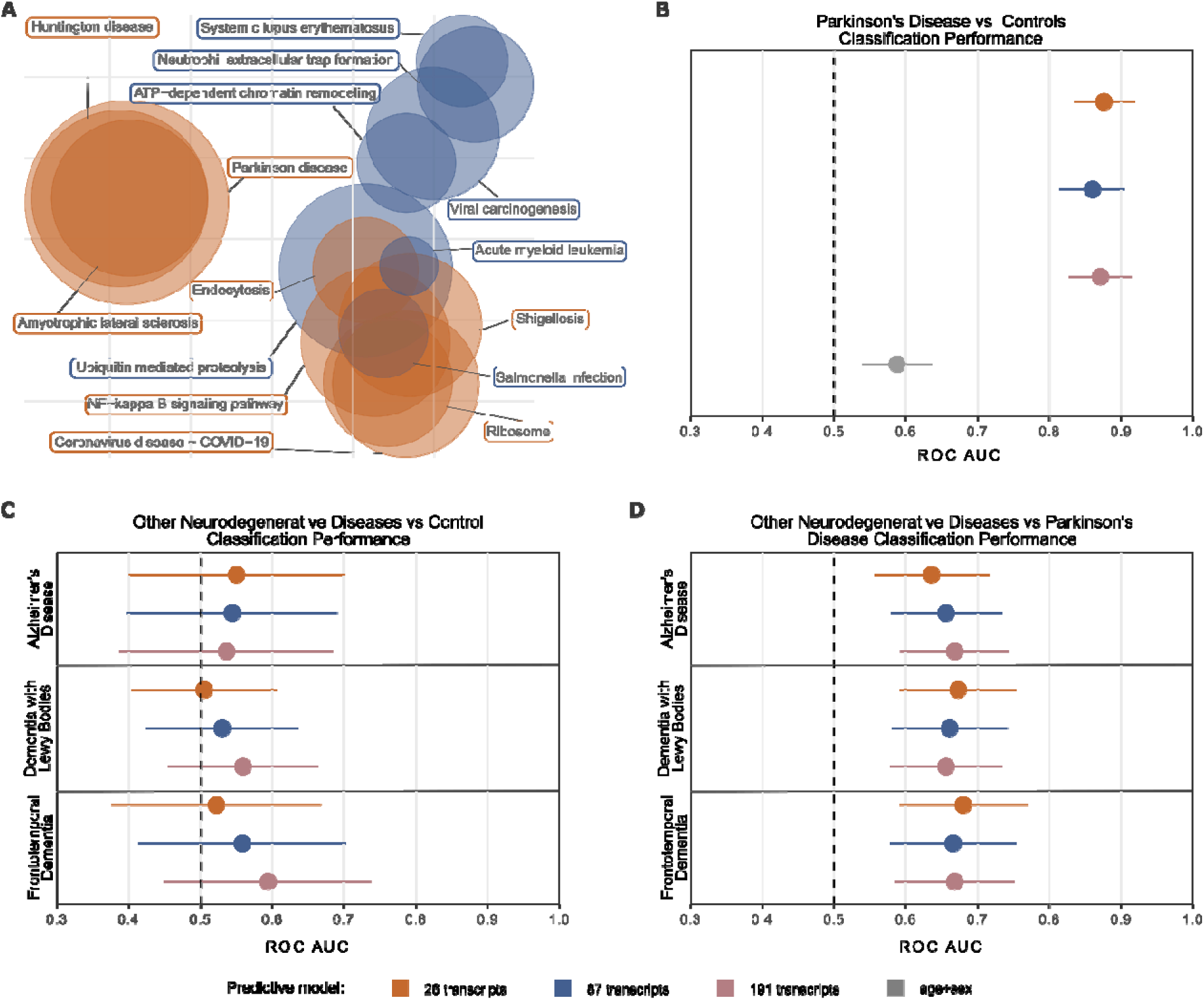
Plasma cfRNA capture signatures associated with PD pathology. **A**. Scatter plot summarizing the significant results of KEGG pathway analysis, in gold are pathways first captured in all the predictive models and in blue pathways captured in 87 and 191 transcript models; Whisker plot showing the performance of the evaluated predictive models to differentiate between **B**. PD and HC; **C**. AD, DLB, or FTD, and HC; and **D**. AD, DLB, or FTD, and PD.

To contextualize the role of transcripts included in the predictive models, we checked whether any of the transcripts were enriched in the pathways identified in KEGG pathway enrichment analyses described including all the DE transcripts. We found that our smallest subset, consisting of 26 transcripts, captured transcripts that pertain to PD, ubiquitin mediated proteolysis, HD, and ALS (Figure 3A). The next subset, with 87 transcripts, further captures transcripts involved in endocytosis (Figure 3A). Next, we checked whether proteins translated from any of the transcripts included in the predictive models were differentially accumulated in plasma and found two proteins, BMP6 and PARP1, to be differentially accumulated.

Finally, we explored whether the selected transcripts reflected motor symptom severity, measured by Unified Parkinson’s Disease Rating Scale (UPDRS) Part III (UPDRS-III), and cognitive status measured by The Montreal Cognitive Assessment (MoCA). UPDRS-III scores were available for both datasets, but not for all participants. Due to sample size (n_HUMT_=78, n_WUSM_=28), we combined HUMT and WUSM data for UPDRS-III analysis and observed that three (*PLAC8*, p=1.82×10^-4^; *PTK2B*, p=0.02; *ATP5F1B*, p=0.04) of the 191 selected transcripts correlated negatively, and one (*RPS27P8*, p=0.02) correlated positively, with UPDRS-III (Supplementary Table 10). MoCA information was available for a subset of the WUSM population (n=28). We found two cfRNA transcripts (*FCGR3A*, p=0.01; *RERE*, p=0.04) with significant positive correlations to MoCA scores (Supplementary Table 10).

### The predictive models are specific to PD

To assess whether the predictive models were specific to PD, we tested the models in samples from Dementia with Lewy bodies (DLB, n=29), Alzheimer’s disease (AD, n=44), and Frontotemporal dementia (FTD, n=16), using two approaches. Firstly, we evaluated if the models could differentiate between each neurodegenerative disease and healthy controls and secondly, if they could discern between PD and DLB, AD, or FTD. The models exhibited low predictive power to differentiate between healthy controls and AD (0.53<AUC<0.55), DLB (0.51<AUC<0.57), or FTD (0.52<AUC<0.59; Figure 3C, Supplementary Table 11), confirming that the models are specific to PD. Models performed slightly better in classifying AD (0.64<AUC<0.67), DLB (0.65<AUC<0.67), or FTD (0.67<AUC<0.68; Figure 3D, Supplementary Table 11) when compared to PD. Given the know association between APOE genotype and AD risk^60^, we tested whether the addition of *APOE* genotype affects the ability of the predictive models to differentiate between AD and PD. With the inclusion of *APOE* information, we observed an increase in the power of our predictive models to discern between AD and PD (0.85<AUC<0.88).

To gain more insight about whether any of the transcripts included in the predictive models were commonly dysregulated across different NDDs, we investigated plasma expression patterns across PD, DLB, AD and FTD. We observe the most remarkable difference in FTD, where the same transcripts seem to be dysregulated in opposite direction to PD (Supplementary Figure 3).

## DISCUSSION

In the present study, we leveraged two independent datasets to identify plasma cfRNAs that were dysregulated in PD participants and, for the first time in PD, employ plasma cfRNAs to develop predictive models that can distinguish between PD and healthy controls. Overall, we identified 2,188 transcripts that were DE in plasma of PD participants. Through pathway analyses we showed that the identified transcripts are part of PD-associated pathways, such as endocytosis^61^, ubiquitin mediated proteolysis^62–64^, PD, and PD-related cellular components and biological processes such as ubiquitin ligase complex^62–64^, primary lysosome, lysosome transport^65^ and exocytosis.^66,67^ Furthermore, we show that our findings in plasma are consistent with those in blood by utilizing publicly available PD blood bulk RNAseq data to replicate 424 of our findings. Additionally, we find that protein products of twelve of the identified acellular transcripts are also dysregulated in plasma. One of the twelve transcripts, COL6A3, has previously been indicated in other neurologic disorders, namely muscular dystrophy^68^ and dystonia^69^. Interestingly, three of the twelve transcripts, *ITGAM*^70^, *SERPINB8*^71^, and *SLC27A4*^72,73^, are associated with dry, flaky skin, which is a common symptom of PD.

Leveraging an *in-house* brain bulk RNAseq dataset, we showed that plasma cfRNA captures changes occurring in the brains of PD participants, most likely due to blood-brain barrier (BBB) leakage.^74,75^ Moreover, we found a significant overlap between transcripts DE in plasma and the corresponding proteins in CSF. Specifically, 24 differentially accumulated CSF proteins are produced from transcripts DE in both plasma and brain. Several of these proteins/transcripts are associated with other neurodegenerative or movement disorders, such as AD (*KLC1*^76^) and dystonia (*SYNE2*^77^), or PD-related pathologies and pathways (*ARF3*^78,79^, *DCTN2*^80^, *HERC1*). Specifically, *ARF3* contributes to the disruption of Golgi apparatus^78,79^ and there is evidence of Golgi fragmentation in PD.^81^ *DCTN2* colocalizes with phosphorylated *SNCA* in Lewy bodies in participants with PD and DLB.^80^ *HERC1* is an E3 ubiquitin protein ligase whose dysregulation leads to alterations in the endosomal system^82^, and both atypical ubiquitination and dysfunction of endosomal system are hallmarks of PD pathobiology.^63^

Lastly, we showed that 196 of the 2188 identified transcripts originate from genomic regions associated with PD. Of those, 121 have some level of support as the driving gene for a given locus.^49,50^ Remarkably, six of these transcripts are encoded by genes nominated by the PD GWAS studies (*ADORA2B, DYRK1A, GCH1, PNA1, MCCC1, TMEM163*), adding additional evidence to the already prioritized genes.^49,83^ All but ADORA2B had high supporting scores, ranging from seven (*MCCC1*) to ten (*DYRK1A*), in the *GWAS* browser. Interestingly, DYRK1A has the same level of support as *SNCA*, a well-known PD-associated gene, in the locus browser^50^, adding evidence of the involvement of *DYRK1A* in the pathobiology of PD.

To date, there are no established biomarkers for accurate PD diagnosis, yet with α-synuclein seed amplification assays (SAA) having promising performance in CSF.^14^ We utilized the identified DE transcripts to develop minimally-invasive predictive models capable of differentiating between PD and healthy controls. We developed three scalable models with high predictive power to classify PD, with AUC values between 0.86 and 0.88. Two of the predictive transcripts, *BMP6* and *PARP1*, are also dysregulated on a protein level in plasma, which opens additional possibilities to leverage the proteins as biomarkers. Interestingly, increased levels of *BMP6*^84^ have been associated with AD, while PARP1 is connected with α-synuclein pathology and PD^85^, but they have not been explored as biomarkers. Regardless, we showed that the transcriptomic models are specific to PD, as they are unable to differentiate between DLB, FTD or AD and healthy controls and capture pathways relevant to the known pathobiology of PD. Notably, we observed that the predictive transcripts are dysregulated in FTD in the opposite direction to PD, suggesting that underlying molecular pathways could be shared between PD and FTD, though regulated differently. Due to the complexity of PD, and the biological interplay between DNA, RNA, and protein functions, we believe that future PD biomarkers might benefit from integrating proteomic and transcriptomic data. Mapping the predictive model transcripts to the pathway analysis results, we show that model transcripts are involved in PD relevant pathways, such as PD, ubiquitin mediated proteolysis, as well as other nervous system disorders, like HD and ALS, adding further evidence of the potential translatability of these models, and their biological relevance.

Four transcripts, *PLAC8, PTK2B, RPS27P8, ATP5F1B*, included in the models correlate significantly with motor symptoms measured by UPDRS-III scale, several of which have known links to PD. *PTK2B* is highly expressed in the nervous system and has been indicated in AD for its role in synaptic homeostasis.^87–89^ In PD, PTK2B is associated with variant rs11060180 (p=1.12×10^−4^), considered to be a PD-risk allele.^90^ Similarly, *ATP5F1B* has no known correlation to PD, but is associated with dystonia, which, like PD, is a movement disorder, further supporting its potential relation with UPDRS-III.^92^ Finally, two transcripts, *RERE* and *FCGR3A*, correlate with dementia symptoms measured by MoCA scores. The latter of the two have already been associated with memory disorders, highlighting the biological relevance of our predictive models.^93^ Together, this further shows that acellular transcripts are truly capturing PD pathology and the complexity of the movement disorder diseases and potentially points to different mechanisms causing the motor versus cognitive symptoms in PD participants.

This study has several limitations. While we accessed two independent populations from different repositories, we are limited by sample size and in the amount of clinical information we had available for HUMT population. This has made it challenging to account for all relevant biological, medical, and technical variables. Further, samples have been stored in the freezer for varying amounts of time, which is known to affect the quality of RNA, and thus can potentially impact our findings. To minimize the effects of missing clinical information and storage time, we removed from the analyses any transcripts that showed selective degradation or were associated with medication in WUSTL population as previously described.^33^ Finally, larger sample sizes of both PD samples, as well as other NDDs, would allow for further validation and sensitivity testing of the developed predictive models.

Nonetheless, this study is the first of its kind and shows that cfRNAs have a potential to aid in diagnosis of PD as cost effective, minimally invasive biomarkers. We identified several plasma transcripts that have already been associated to PD or relevant pathways, correlate with symptom severity, and could potentially be leveraged for disease monitoring. On top of that not only are some of those transcripts dysregulated in different tissues, but they are also encoded in known PD loci. Additionally, some of them result in alterations in protein abundance when compared to healthy controls in relevant tissues. Overall, we believe that we have demonstrated that cfRNA has the power to capture changes relevant to PD pathology, has the potential to be translated to the clinic, and if replicated and validated in larger samples sizes, would benefit the whole PD community by providing non-invasive biomarkers, that are cost-effective and can be implemented in remote areas, providing access to care for all.

## DATA AVAILABILITY

Data is available from the NIAGADS Data Sharing Service (NIAGADS dss), accession number NG00142. All original code has been deposited at GitHub (https://github.com/Ibanez-Lab/PlasmaCellFreeRNA-ParkinsonDisease) and is publicly available as of the date of publication. Any additional information required to reanalyze the data reported in this paper is available from the lead contact upon request.

## ACKNOWLEDGEMENTS

We thank all the participants and their families along with the institutions and all the staff who provided plasma tissue, without whom this study would not have been possible. This work was supported by access to equipment made possible by the Hope Center for Neurological Disorders, the Neurogenomics and Informatics Center (NGI: https://neurogenomics.wustl.edu/) and the Departments of Neurology and Psychiatry at Washington University School of Medicine. This work was supported by grants from the National Institutes of Health K99/R00-AG062723, P01AG003991, RO1 NS075321, P30 AG066444, P01 AG03991, and P01 AG026276, the Michael J Fox Foundation (MJFF-021599 and MJFF-025292), the Alzheimer’s Drug Discovery Foundation (GDAPB-201807-2015632), Bright Focus Foundation (A2021033S), and the Department of Defense (W81XWH2010849).

## AUTHOR CONTRIBUTIONS

LI, and AB conceived and wrote this article. LI conceptualized and designed the research plan. LI and AB designed the analysis plan. AB, ACG, CM, RK, IAD, KB, and IS processed all the data and performed the analyses. IA and SY obtained and processed the samples. IA and ML obtained and curated clinical information. AB, ACG, CM, RK, IAD, KB, IS, SY, JS, DW, ML, IA, JP, SN, PP, GZ, JB, and LI discussed the project, revised the manuscript, and provided critical feedback.

