## SupplementaryFigures for "Plasma acellular transcriptome contains Parkinson’s disease signatures that can inform clinical diagnosis"

Supplementary Figures

**Supplementary Figure 1.** Volcano plot summarizing meta-analyses results. The Y-axis represents the -log_10_ of the meta-analysis p-value, and the X-axis is the weighted mean of the log_2_ fold change from the analysis of the individual populations, HUMT and WUSM. Significantly DE transcripts are highlighted in orange.

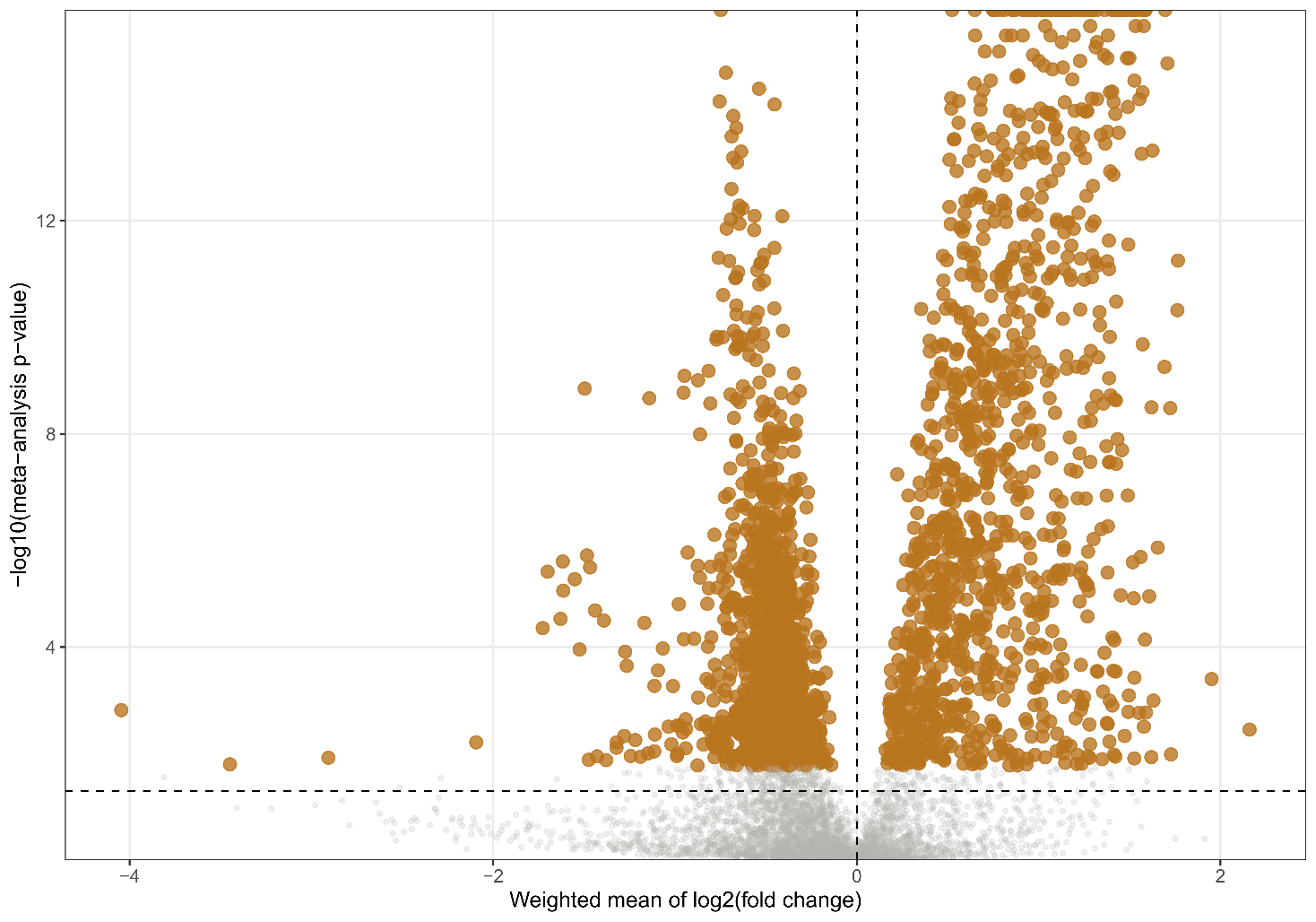

**Supplementary Figure 2. Feature Selection.** Model performance in the training population for the increasing KLD value cutoffs. The Y-axis represents the AUC of each model, X-axis represents the KLD value used as threshold, and in orange are highlighted the models we selected for further analyses.

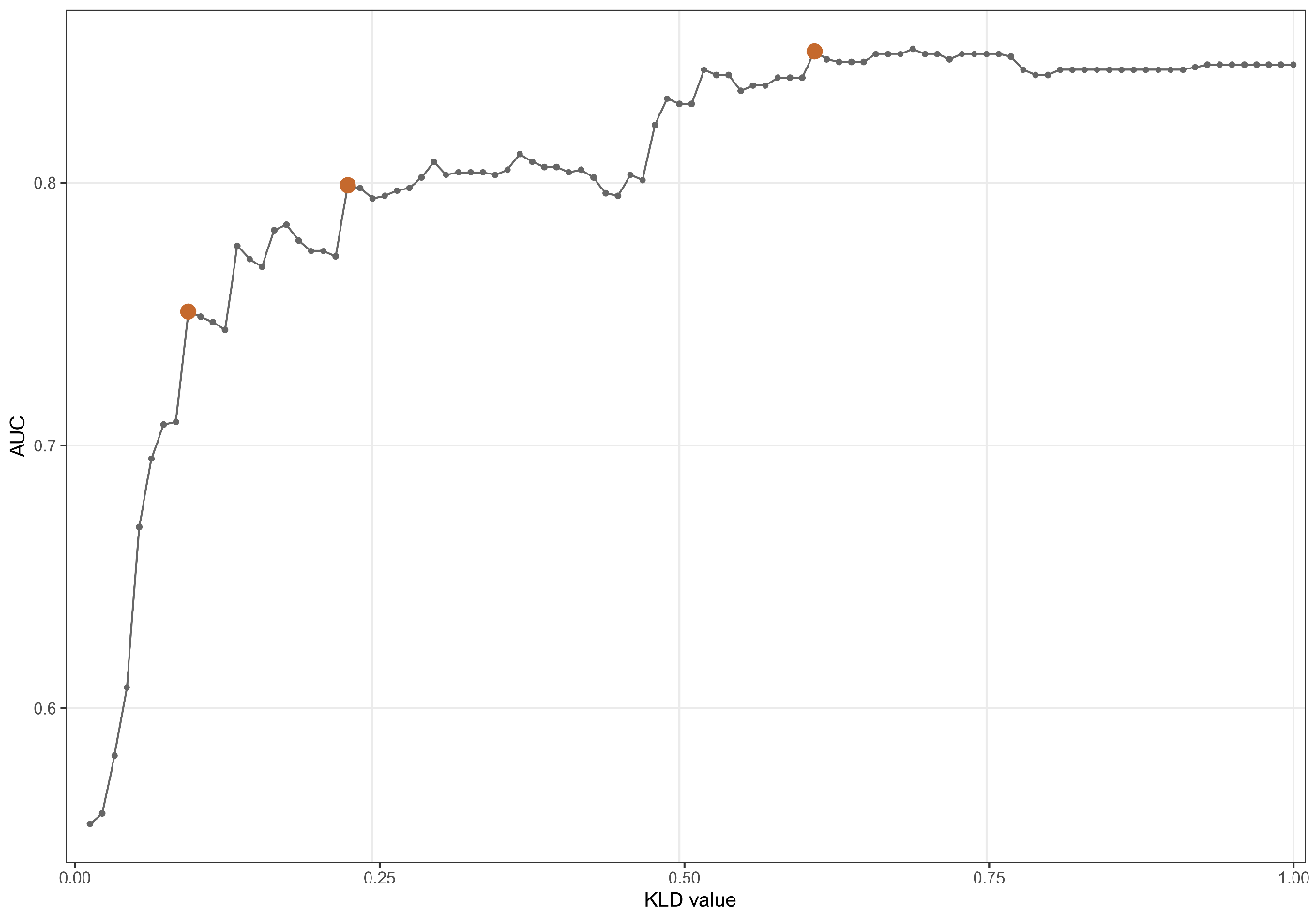

**Supplementary Figure 3. Heatmap** comparing the log_2_ fold change for the differential expression of the 214 transcripts included in the predictive models in each neurodegenerative disease. Transcripts are sorted in decreasing order of the log_2_ fold change in the comparison between PD and healthy controls.

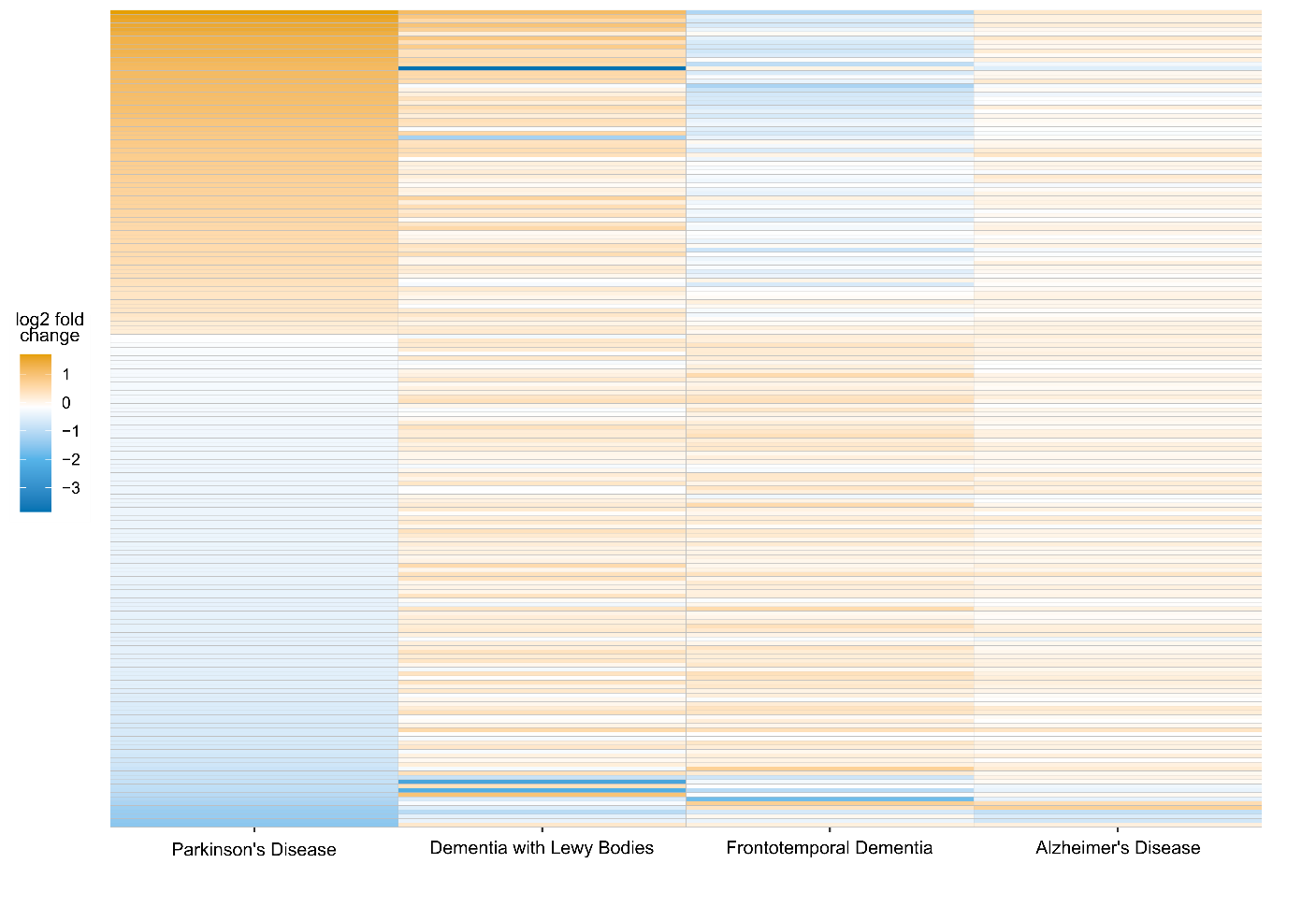
